# Direct Oral Factor-Xa Inhibitors in Patients with Acute Venous Thromboembolism and Renal Impairment. A randomized controlled trial

**DOI:** 10.64898/2026.08.03.26359634

**Authors:** Patrick Mismetti, Laurent Bertoletti, Antoine Elias, Carine Assante, Olivier Sanchez, Jeannot Schmidt, Emilie Presles, Céline Chapelle, Sandrine Accassat, Francis Couturaud, Isabelle Mahé, Silvy Laporte, VERDICT Investigators

## Abstract

**BACKGROUND:** In patients with venous thromboembolism (VTE), renal impairment increases the risks for both recurrence and bleeding. Because these patients are underrepresented in clinical trials, we assessed whether standard direct factor Xa inhibitor (DXI) lead-in followed by early dose reduction was noninferior to standard anticoagulation in patients with acute VTE and moderate-to-severe renal impairment.

**METHODS:** The VERDICT trial was a randomized, prospective, multicenter, open-label, blinded-endpoint, noninferiority trial. Consecutive patients with acute proximal deep-vein thrombosis or pulmonary embolism and chronic renal impairment (creatinine clearance 15–50 mL/min) were randomized 1:1 to an early DXI dose reduction strategy or standard therapy (heparin plus a vitamin K antagonist). Patients allocated to the DXI strategy underwent a second 1:1 randomization to apixaban or rivaroxaban, each administered at an initial standard lead-in dose followed by early dose reduction. The primary outcome was net clinical benefit at 3 months, defined as the composite of major bleeding and symptomatic recurrent VTE.

**RESULTS:** Due to slow recruitment, the trial was prematurely terminated after enrolling 200 of the planned 800 patients (DXI: n=104; standard: n=96). The median age was 85.9 years, 31% were male, and 29.0% had severe renal impairment. The primary outcome occurred in 8 patients (7.7%) in the DXI group and 9 patients (9.3%) in the standard therapy group (adjusted subhazard ratio [sHR], 0.87; 95% CI, 0.26 to 2.87; P = 0.19 for noninferiority; noninferiority margin 1.30). Major bleeding occurred in 6.7% and 6.2% of patients and recurrent VTE occurred in 1.0% and 3.1% of patients, respectively.

**CONCLUSION:** In patients with VTE and moderate-to-severe renal impairment, noninferiority of an early DXI dose-reduction strategy versus standard therapy could not be demonstrated for net clinical benefit. Although no major differences in efficacy or safety outcomes were observed between groups, the reduced sample size precludes definitive conclusions.

**Clinical Trial Registration:** URL: https://clinicaltrials.gov. Unique identifier: NCT02664155

**Clinical Perspective:** *What Is New?:*

- The VERDICT trial is the first randomized controlled trial specifically designed to assess an initial frontloaded standard dose of oral direct factor Xa inhibitor followed by early dose reduction strategy versus standard anticoagulation therapy for acute venous thromboembolism in patients with moderate-to-severe renal impairment.
- Reflecting real-world clinical practice, the study successfully enrolled an exceptionally elderly and frail population with a median age of 85.9 years.
- A meta-analysis incorporating VERDICT and renal impairment subgroups from previous VTE trials showed a 35% relative reduction in net clinical benefit and a 53% relative reduction in major bleeding with DXI versus standard therapy with heparins/VKA.

*What Are the Clinical Implications?:*

- Although premature trial termination limited statistical power to formally demonstrate noninferiority, the findings did not differ from those observed in renal impairment subgroups of previous randomized VTE trials.
- The available evidence provides a rationale for evaluating an early dose-reduction strategy in frail patients with VTE.

## INTRODUCTION

Venous thromboembolism (VTE), which includes deep-vein thrombosis (DVT) and pulmonary embolism (PE), is a common and potentially life-threatening condition worldwide. Effective anticoagulant therapy is essential to prevent thrombotic recurrence and reduce mortality in affected patients. The management of VTE has evolved considerably over the past decades, driven by advances in anticoagulant pharmacology and an improved understanding of patient-specific risk profiles [1].

Standard anticoagulant therapy—consisting of initial treatment with low-molecular-weight heparin (LMWH) or unfractionated heparin (UFH), followed by a vitamin K antagonist (VKA)—has long been the mainstay of VTE management, but has practical and clinical limitations. Indeed, heparin-based regimens can be cumbersome, particularly in frail or hospitalized patients, and are associated with specific risks such as heparin-induced thrombocytopenia. Moreover, VKAs require regular laboratory monitoring, have a narrow therapeutic window, and are subject to numerous drug interactions.

Over the past decade, direct oral anticoagulants (DOACs), particularly direct anti-Xa inhibitors (DXIs) such as apixaban and rivaroxaban, have transformed VTE management. Their predictable pharmacokinetics, fixed dosing, and lack of routine monitoring have made them the first-line therapy for most patients. Pivotal phase III trials have demonstrated that direct oral anticoagulants are at least as effective as heparin/VKA regimens in preventing recurrent VTE, with lower rates of major bleeding events [2–5].

However, these trials predominantly enrolled relatively young patients (around 55 years old) with preserved renal function. Impaired renal clearance alters the pharmacokinetics of most anticoagulants and can increase the risk of hemorrhagic complications. Yet, renal impairment is present in more than 20% of patients presenting with VTE [6] and is associated with a markedly increased risk of both recurrent VTE and major bleeding [7,8]. Patients with severe renal impairment (creatinine clearance estimated by Cockcroft and Gault formula < 30 ml/min /1.73m2) were systematically excluded from pivotal studies and those with moderate renal impairment (creatinine clearance 30–50 ml/min) were markedly underrepresented [9], less than 6% of the study populations [10]. As a result, comparative data evaluating anticoagulation strategies in patients with moderate to severe renal impairment are lacking, and the optimal therapeutic approach in this high-risk population remains uncertain.

Among direct oral anticoagulants, apixaban and rivaroxaban are unique in allowing a single-drug regimen from the acute phase onward, without requiring initial heparin therapy. While a front-loaded treatment dose is required during the acute phase of VTE, subsequent dose reduction in patients with renal impairment may optimize the net clinical benefit by limiting drug accumulation and bleeding while maintaining antithrombotic efficacy.

In this context, renal function is traditionally estimated using the Cockcroft-Gault formula, which was systematically used to determine trial eligibility in pivotal phase III VTE studies. However, while a creatinine clearance cut-off of 50 mL/min is widely used for dose reduction in atrial fibrillation, patients with moderate renal impairment (30-50 mL/min) received full-dose regimens in the landmark VTE trials. Given that the Cockcroft-Gault formula frequently under- or overestimates clearance in many subgroups, the optimal approach to balancing thrombotic and bleeding risks in these patients remains unestablished.

The objective of this study was therefore to assess whether an initial standard front-loaded DXI regimen (apixaban 10 mg bid for 7 days or rivaroxaban 15 mg bid for 21 days) followed by early dose reduction (apixaban 2.5 mg bid or rivaroxaban 15 mg od) was noninferior to standard therapy (LMWH or UFH followed by VKA) with respect to net clinical benefit in patients with acute symptomatic VTE and moderate to severe chronic renal impairment.

## METHODS

### Trial Design and Oversight

VERDICT is a French, prospective, randomized, multicenter, open-label, noninferiority study with blinded adjudication of outcome events, comparing initial frontloaded standard dose of DXI followed by early dose reduction with standard therapy. The trial protocol was approved by the Ethics Committee (CPP Sud-Est I) on 11 April 2016 and by the French National Agency for the Safety of Medicines and Health Products (ANSM) on 13 May 2016. The steering committee was responsible for the design and oversight of the trial, the development of the protocol, the analysis of the data, the writing of the manuscript, and the decision to submit the manuscript for publication. The investigators gathered the data.

The trial was managed by the University Hospital of Saint-Étienne, France, and supported by a grant from the French Ministry of Health through the National Hospital Clinical Research Program (PHRC-N-15-651). The trial was registered with ClinicalTrials.gov, NCT02664155. It was coordinated by the Clinical Research Unit of the University Hospital of Saint-Étienne and the steering committee. Data were collected and maintained using the EnnovClinical electronic data capture system and analyzed by the Clinical Research Unit of the University Hospital of Saint-Étienne. A central adjudication committee, whose members were unaware of the treatment assignments, reviewed all suspected outcome events and causes of death. An independent data and safety monitoring board periodically reviewed trial safety. The trial investigators and committees are listed in the Supplementary Appendix. The trial was conducted and reported in accordance with CONSORT guidelines (Supplementary Table 1).

### Patients

Patients with moderate (creatinine clearance 30–50 ml/min) or severe (creatinine clearance 15– 29 ml/min) renal impairment and acute symptomatic VTE—defined as proximal DVT of the lower limb or PE objectively confirmed within 72 hours—were eligible to participate if they were scheduled to receive anticoagulant treatment for at least 3 months. Detailed inclusion and exclusion criteria are provided in the Supplementary Appendix.

### Randomization and Trial Intervention

This was a two-stage randomization design. Patients were first randomly assigned, in a 1:1 ratio, to either the DXIs group or the standard therapy group (UFH or LMWH plus), with treatment administered for at least 3 months. Participants assigned to the DXIs group were subsequently randomized to receive either apixaban 10 mg twice daily for 7 days directly followed by 2.5 mg twice daily for 3 months, or rivaroxaban 15 mg twice daily for 21 days directly followed by 15 mg once daily for 3 months.

In the standard therapy group, anticoagulation was adapted to the severity of renal impairment: patients with moderate renal impairment received subcutaneous LMWH adjusted to body weight for at least 5 days in combination with VKA (INR 2-3); those with severe renal impairment received either subcutaneous or intravenous UFH, or subcutaneous LMWH (adjusted to body weight) for at least 5 days, in combination with VKA (INR 2-3). Further details on treatment regimens and design are provided in the Supplementary Appendix. Randomization was performed centrally using an interactive web-response system. The allocation sequence was computer-generated using permuted blocks of varying sizes. Randomization was stratified according to trial center and severity of renal impairment (moderate or severe).

### Outcome Measures

The primary outcome was net clinical benefit, defined as the composite of centrally adjudicated symptomatic fatal or nonfatal recurrent VTE and adjudicated major bleeding events, assessed over 3-months. In a hierarchical procedure, the key secondary outcomes were, first, major bleeding and, second, symptomatic recurrent VTE over 3-months. Other safety outcomes included death from any cause, clinically relevant bleeding (composite of major or clinically relevant nonmajor bleeding), and symptomatic cardiovascular events. Detailed outcome definitions are provided in the Supplementary Appendix.

### Statistical Analysis

A prespecified hierarchical testing strategy was used to assess the noninferiority of DXIs compared with standard therapy in the following order: (1) net clinical benefit, (2) major bleeding, and (3) recurrent VTE. Noninferiority for the primary outcome was assessed using a prespecified margin of 1.30 for the upper boundary of the two-sided 95% confidence interval of the subdistribution hazard ratio (subHR). The rationale for the noninferiority margin is provided in the Supplementary Appendix.

The composite primary endpoint combined recurrent VTE and major bleeding to assess net clinical benefit. Assuming event rates of 6% in the DXIs group and 9% in the standard therapy group for the primary outcome, 800 patients were required to provide 85% power to demonstrate noninferiority at a one-sided type I error of 2.5%, allowing for 5% loss to follow-up. Details of the sample size calculation, including assumptions for the key secondary outcomes, are provided in the Supplementary Appendix. The trial was prematurely terminated by the Steering Committee because of slow recruitment related to the COVID-19 pandemic, independently of any efficacy or safety assessment, after 200 of the planned 800 patients had been enrolled.

Time-to-event analyses were performed using Fine and Gray regression models accounting for the competing risk of death, with the randomization strata as covariates [11,12]. Noninferiority for the primary outcome was assessed in both the intention-to-treat and per-protocol populations. If demonstrated in both populations, hierarchical testing proceeded to major bleeding and subsequently to recurrent VTE. The intention-to-treat population included all randomized patients. The per-protocol population included patients who received at least one dose of study treatment and had no major protocol violations (Supplementary Appendix).

Treatment effects were expressed as subRHs with two-sided 95% confidence intervals. Follow-up was censored at the last known contact for patients without an event. Missing data were not imputed. Prespecified subgroup analyses were performed for the primary and key secondary outcomes. Analyses were performed using SAS version 9.4 (SAS Institute).

### Meta-analysis

A meta-analysis was performed using results on net clinical benefit at the end of the treatment period (3 to 12 months), recurrent VTE, and major bleeding from four previously randomized controlled trials (RCTs) comparing DXIs with LMWHs/VKA in patients with VTE [13–15,9]. Available subgroup data for patients with moderate renal impairment (creatinine clearance 30– 50 mL/min), together with the data reported here, were included in the analysis. We used a fixed-effects model based on the logarithm of the risk ratio (RR), weighted by the inverse of the variance, to combine results from the individual trials. Statistical heterogeneity among studies was assessed using Cochran’s Q statistic, and consistency across studies was quantified using the I^2^ statistic [16]. The meta-analysis was performed using R software (version 4.5.2; meta package, available at www.r-project.org).

## RESULTS

### Patients and Treatments

From October 19, 2016 to November 30, 2021 a total of 200 patients were enrolled at 24 centers in France and underwent randomization; 104 were assigned to the DXIs group and 96 to the standard therapy group (Figure 1). The median follow-up duration was 6.0 months (interquartile range, 5.8 to 6.3), and 191 of 200 patients (95.5%) were followed at least 3 months. Baseline demographic and clinical characteristics were similar in the two groups (Table 1). The median age was 85.9 years (interquartile range [IQR], 80.8 to 90.1), 62 patients (31.0%) were men, and the median body weight was 68.7 kg (IQR, 59.0 to 78.0). The index event was PE in 166 patients (83.0%) and isolated lower limb DVT in 33 patients (16.5%). Overall, 142 patients (71.0%) had moderate renal impairment and 58 (29.0%) had severe renal impairment. Of the 104 patients assigned to the DXIs group, 1 (1.0%) did not receive the study drug; 54 (52.4%) received apixaban and 49 (47.6%) received rivaroxaban. In the standard therapy group, 67 patients (69.8%) received LMWH plus VKA, and 29 (30.2%) received UFH plus VKA.

**Figure 1.**
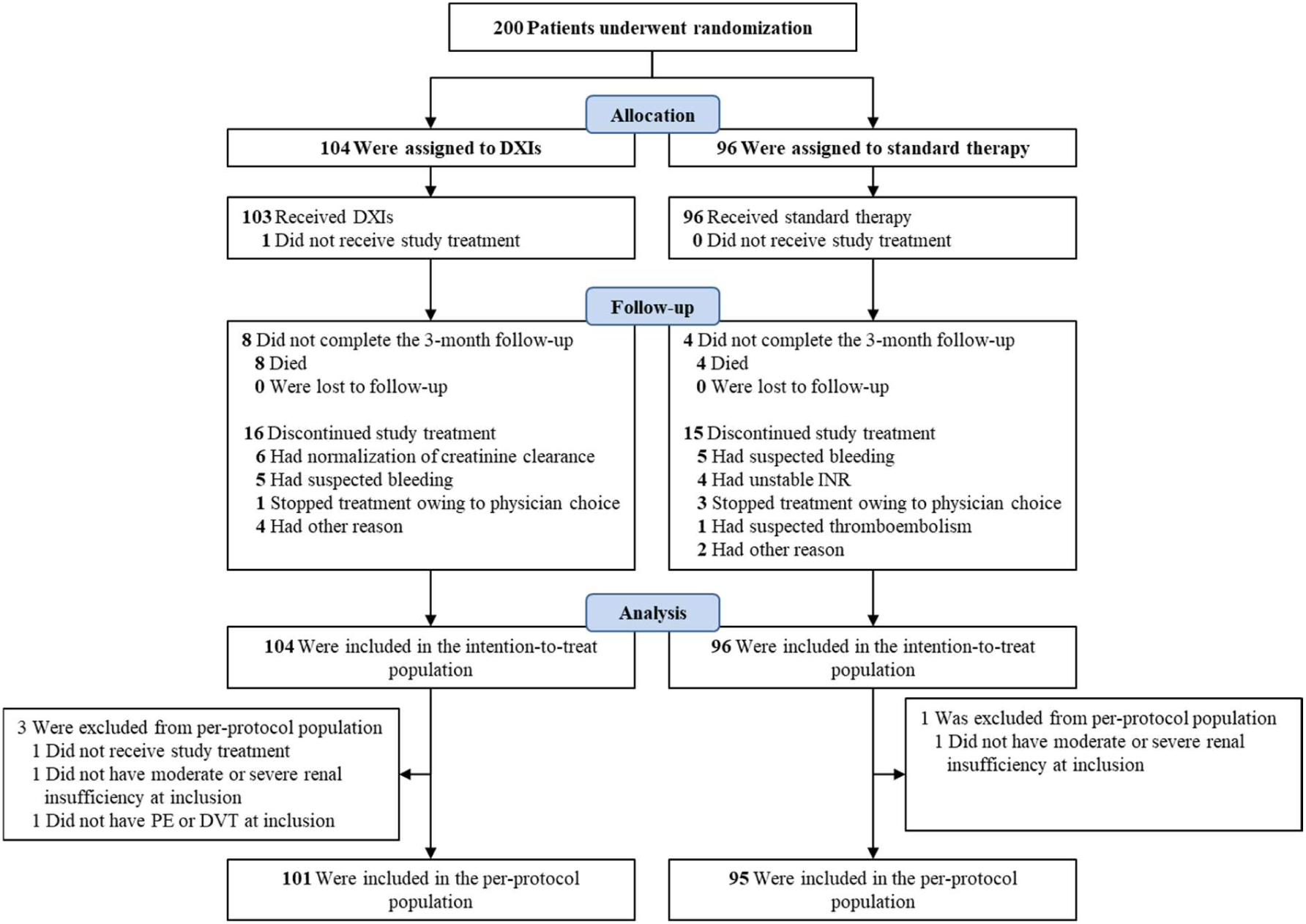
Randomization, Treatment, and Follow-up of the Patients.

**Table 1.** Demographic and Clinical Characteristics at Baseline.

| **Characteristic\*** | **DXIs**<br>**(N=104)** | **Standard**<br>**therapy**<br>**(N=96)** |
| --- | --- | --- |
| Age – years | 84.6±9.7 | 84.1±8.2 |
| Age ≥ 75 years – no. (%) | 90 (86.5) | 83 (86.5) |
| Age ≥ 90 years – no. (%) | 28 (26.9) | 23 (24.0) |
| Male sex – no. (%) | 31 (29.8) | 31 (32.3) |
| Body weight – kg | 68.1±15.0 | 68.8±13.7 |
| Body weight < 50 kg - no. (%) | 11 (10.6) | 8 (8.3) |
| Body mass index – kg/m<sup>2</sup> | 25.9±5.0 | 26.1±5.1 |
| Creatinemia – μmol/l | 129.0±55.5 | 130.9±53.1 |
| Creatinine clearance** – ml/min | 34.7±8.4 | 34.7±7.9 |
| Renal impairment<sup>#</sup> – no. (%) |  |  |
| Moderate | 74 (71.2) | 68 (70.8) |
| Severe | 30 (28.8) | 28 (29.2) |
| Qualifying diagnosis of VTE – no. (%) |  |  |
| PE with or without DVT | 85 (81.7) | 81 (84.4) |
| Isolated proximal DVT | 18 (17.3) | 15 (15.6) |
| None | 1 (1.0) | 0 (0.0) |
| Major transient risk factors in the past 3 months – no. (%) |  |  |
| Surgery with general anesthesia lasting > 30 minutes | 2 (1.9) | 6 (6.3) |
| Hospitalization with bed-rest > 3 days | 11 (10.6) | 9 (9.4) |
| Minor transient risk factors in the past 2 months – no. (%) |  |  |
| Fracture or immobilization of a lower limb | 0 (0.0) | 1 (1.0) |
| Bed-rest ≥ 3 days for acute medical disease | 5 (4.8) | 5 (5.2) |
| Persistent risk factors – no. (%) |  |  |
| Active cancer | 7 (6.7) | 4 (4.2) |
| Chronic inflammatory bowel or autoimmune pathology | 7 (6.7) | 5 (5.2) |
| History of VTE – no. (%) | 38 (36.5) | 40 (41.7) |
| Chronic pulmonary disease – no. (%) | 18 (17.3) | 19 (19.8) |
| Diabetes – no. (%) | 25 (24.0) | 18 (18.9) |
| Concomitant antiplatelet therapy – no. (%) | 23 (22.1) | 18 (18.8) |

\* Plus-minus values are means ± SD. Percentages may not total 100 because of rounding.
\*\* Creatinine clearance estimated by the Cockcroft and Gault formula

The median duration of treatment in the overall population was 3.0 months (interquartile range, 2.7 to 3.2). Reasons for permanent discontinuation of the study treatment are provided in Table S2.

### Primary outcome and key secondary outcomes

In the intention-to-treat population, net clinical benefit occurred in 8 patients (cumulative incidence, 7.7%) in the DXIs group and in 9 patients (cumulative incidence, 9.3%) in the standard therapy group (adjusted sub-hazard ratio, 0.87; 95% confidence interval [CI], 0.26 to 2.87; P = 0.19 for noninferiority; prespecified noninferiority margin, 1.30) (Table 2 and Figure 2). In the per-protocol population, the adjusted subdistribution hazard ratio was 0.75 (95% CI, 0.21–2.65).

**Figure 2.**
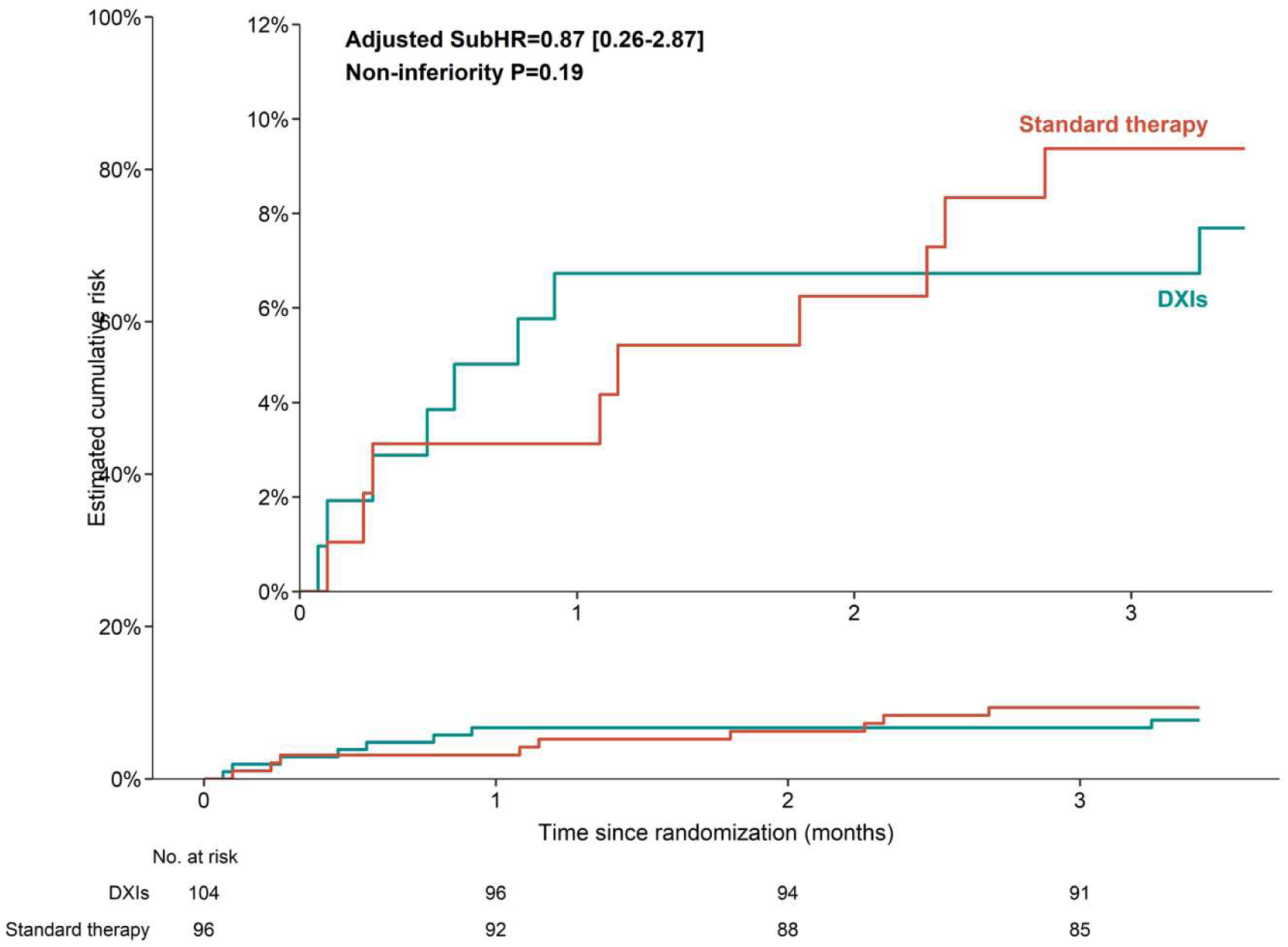
Net clinical benefit (Intention-to-Treat Population). Shown is the cumulative incidence of net clinical benefit (primary efficacy outcome defined as a composite of recurrent symptomatic venous thromboembolism or major bleeding) among patients who received direct oral anticoagulants (DXIs group) or standard therapy (UFH/LMWH followed by VKA). The P value was for noninferiority (margin for the upper boundary of the 95% confidence interval of the subdistribution hazard ratio [subhazard ratio], 1.30). The inset shows the same data on an expanded Y axis.

**Table 2.** Clinical Outcomes during the Trial Period in Intention-to-Treat population.

| **Outcomes at 3 months** | **DXIs**<br>**(N=104)** | **Standard**<br>**therapy**<br>**(N=96)** | **Adjusted**<br>**Subhazard**<br>**Ratio\***<br>**(95% CI)** | **P-value** |
| --- | --- | --- | --- | --- |
| *Number (percent)* |  |  |  |  |
| Primary outcome: net clinical benefit † | 8 (7.7) | 9 (9.3) | 0.87 (0.26-2.87) | 0.19 |
| Key secondary outcomes |  |  |  |  |
| Major bleeding | 7 (6.7) | 6 (6.2) | 1.25 (0.31-4.93) | - |
| Fatal bleeding | 0 (0.0) | 1 (1.0) | - | - |
| Recurrent symptomatic venous thromboembolism‡ | 1 (1.0) | 3 (3.1) | 0.25 (0.02-3.00) | - |
| Pulmonary embolism | 1 (1.0) | 2 (2.1) | - | - |
| Fatal pulmonary embolism | 0 (0.0) | 0 (0.0) | - | - |
| Unexplained sudden death | 1 (1.0) | 0 (0.0) | - | - |
| Proximal deep-vein thrombosis | 0 (0.0) | 2 (2.1) | - | - |
| Other secondary outcomes |  |  |  |  |
| Death from any cause | 8 (7.7) | 4 (4.2) | 1.81 (0.54-6.09) | - |
| Clinically relevant bleeding | 14 (13.4) | 12 (12.4) | 1.10 (0.51-2.36) | - |
| Symptomatic cardiovascular events | 0 (0.0) | 0 (0.0) | - | - |

Percentages are the cumulative incidence and thus may not calculate as expected.
\* adjusted for the randomization strata.
† Composite of recurrent symptomatic venous thromboembolism and major bleeding. The P value is for noninferiority (margin for the upper boundary of the 95% confidence interval, 1.30).
‡ One patient in the standard therapy group had a pulmonary embolism associated with proximal deep-vein thrombosis.

Major bleeding, occurred in 7 patients (cumulative incidence, 6.7%) in the DXIs group and in 6 patients (cumulative incidence, 6.2%) in the standard therapy group (adjusted subhazard ratio, 1.25; 95% CI, 0.31 to 4.93) (Table 2, Figure 3A). Two bleeding episodes involved a critical organ—one intracranial and one retroperitoneal—and one bleeding episode was fatal; all three events occurred in the standard therapy group.

**Figure 3.**
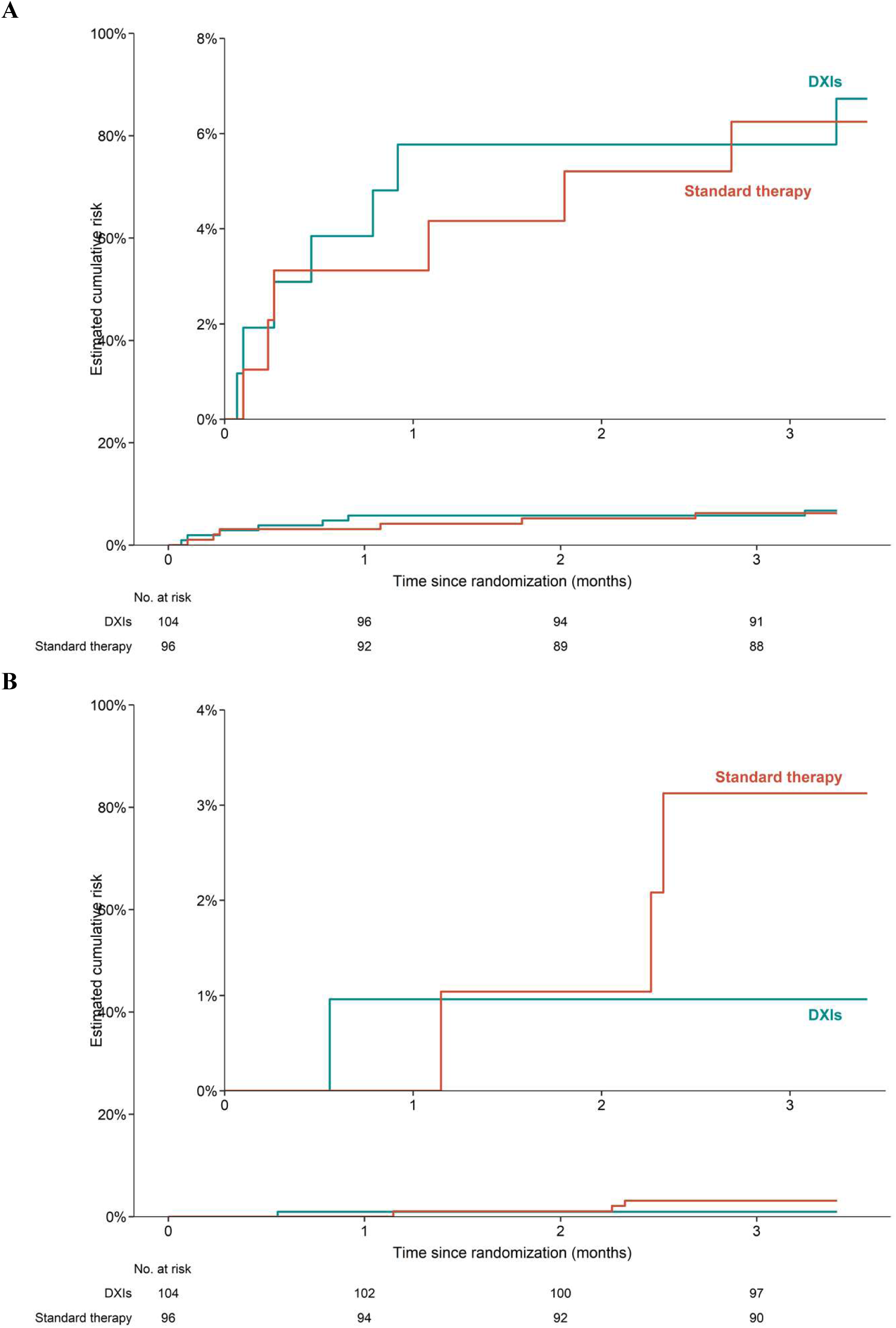
Key secondary outcomes: A) Major bleeding, B) Symptomatic recurrent venous thromboembolism. Shown is the cumulative incidence of A) major bleeding (first key secondary outcome) and B) symptomatic recurrent venous thromboembolism (secondary key secondary outcome) among patients who received direct oral anticoagulants (DXIs group) or standard therapy (UFH/LMWH followed by VKA). The inset shows the same data on an expanded Y axis.

Recurrent symptomatic VTE occurred in 1 patient (cumulative incidence, 1.0%) in the DXIs group and in 3 patients (cumulative incidence, 3.1%) in the standard therapy group (adjusted subhazard ratio, 0.25; 95% CI, 0.02 to 3.00) (Table 2, Figure 3B).

Subgroups analyses for net clinical benefit, major bleeding, and recurrent symptomatic VTE are shown in Figure S1, and S2. Results were similar across the subgroups.

### Secondary Outcomes

Death from any cause occurred in 8 patients (cumulative incidence, 7.7%) in the DXIs group and in 4 patients (cumulative incidence, 4.2%) in the standard therapy group (Table 2 and Table S3). Among patients in the DXIs group, two deaths were attributed to cancer, one was a sudden unexplained death for which PE could not be ruled out, three were due to end-stage heart failure, and two due to general decline. In the standard therapy group, one death was attributed to fatal bleeding, one to acute renal impairment complicated by severe dehydration and septic shock secondary to pneumonia, one was due to end-stage heart failure, and one to severe respiratory failure. No objectively confirmed cases of fatal PE occurred.

Clinically relevant bleeding occurred in 14 patients (cumulative incidence, 13.4%) in the DXIs group and in 12 patients (cumulative incidence, 12.4%) in the standard therapy group (Table 2). No symptomatic cardiovascular events were reported during the study.

### Meta-analysis

The updated meta-analysis pooled data from RCTs—including the four major RCTs and the VERDICT study—encompassing a total of 1,733 patients with VTE and moderate or severe renal impairment (Table S4 and Figure 4). Overall, DXIs were associated with a significantly 35% reduction in the composite net clinical benefit outcome compared with LMWH/VKA (RR 0.65, 95% CI 0.43–0.97). Analysis of the individual components of the composite outcome showed no significant difference between DXIs and LMWHs/VKA for recurrent VTE (RR 0.73, 95% CI 0.44-1.19), although the point estimate favored DXIs, whereas DXIs were associated with a significantly 53% lower risk of major bleeding (RR 0.47, 95% CI 0.25-0.90). The analysis included both conventional- and reduced-dose DXI regimens. Although subgroup analyses were underpowered to formally assess effect modification according to dose and confidence intervals were wide, no obvious qualitative difference in treatment effect was observed between conventional- and reduced-dose DXI regimens.

**Figure 4.**
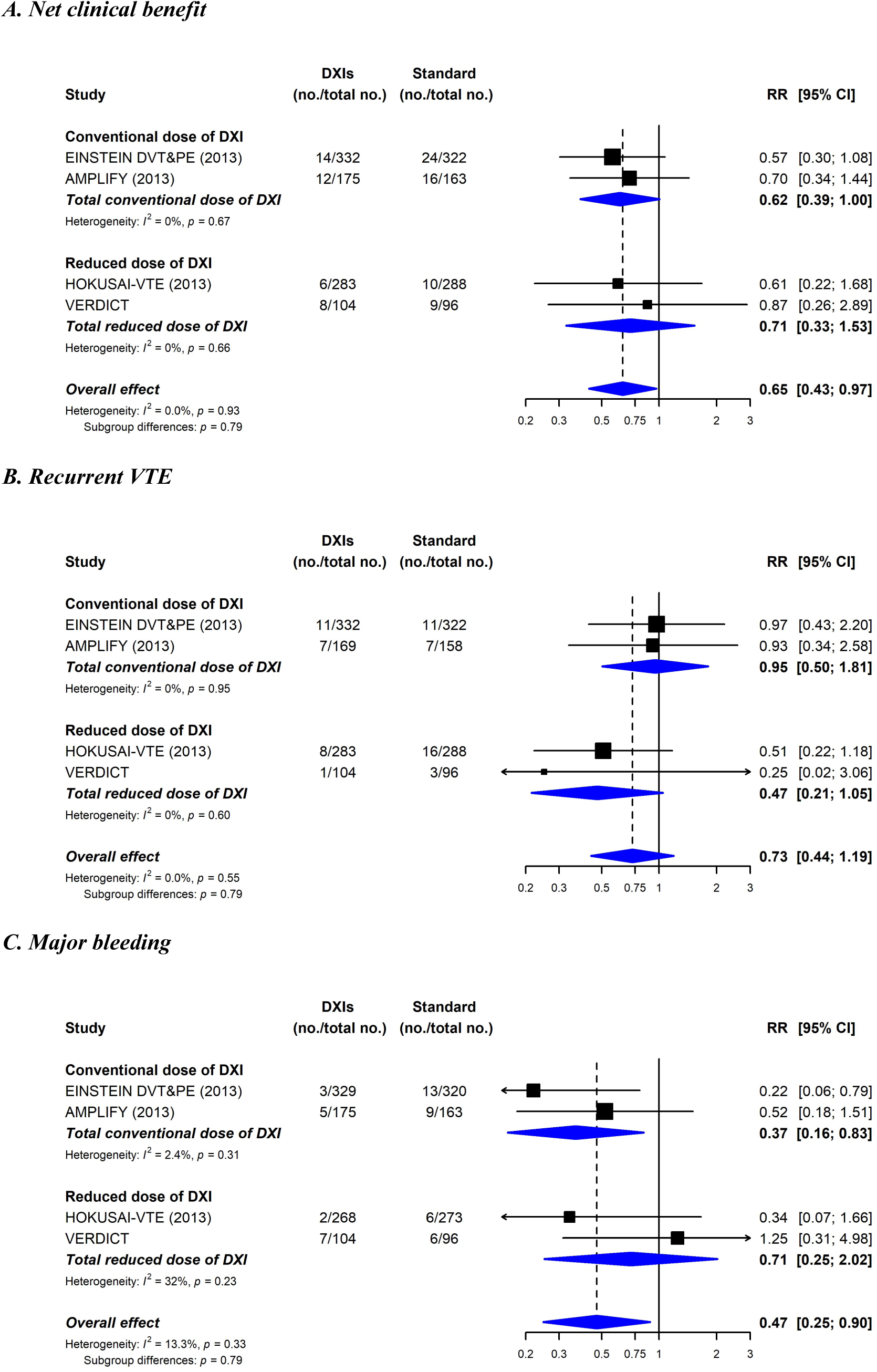
Updated meta-analysis of randomized trials that compared direct oral anticoagulants (DXIs group) or standard therapy (UFH/LMWH followed by VKA) in patients with VTE and moderate or severe renal impairment (creatinine clearance < 50 ml/min). Shown is the meta-analysis of A) net clinical benefit, B) recurrent VTE, and C) major bleeding.

## DISCUSSION

The VERDICT trial did not demonstrate noninferiority of an early dose-reduction of direct factor Xa inhibitor strategy compared with standard anticoagulation for net clinical benefit in patients with VTE and moderate-to-severe renal impairment.

To our knowledge, the VERDICT trial is the first randomized controlled trial specifically designed to evaluate initial frontloaded standard dose followed by early dose reduction of DXIs for acute VTE treatment in patients with impaired renal function. A key methodological feature of VERDICT was the definition of renal impairment using the Cockcroft–Gault formula, consistent with the approach used in historical phase III VTE trials. As a result, the study enrolled an particularly old and frail population (median age > 85 years) with low body weight, markedly different from the younger and lesser comorbid populations included in pivotal VTE trials, and representative of a patient group that is notoriously difficult to screen and enroll in randomized clinical studies.

Although the trial did not demonstrate noninferiority, the observed event rates did not reveal any major efficacy or safety concerns with the DXI strategy used in this trial compared to the standard anticoagulant strategy (LMWH/VKA for 30-50 et UFH/VKA for <30). Given the extreme frailty, advanced age, and high bleeding risk of the enrolled population, maintaining full-dose anticoagulation beyond the initial loading dose treatment phase is questionable. Dose reduction of DXIs according to age, body weight, and renal function is already established in patients with atrial fibrillation [17,18]. Although extrapolation to acute VTE should be made cautiously, the characteristics of the VERDICT population underscore the clinical importance of exploring tailored anticoagulation strategies in this setting. Notably, most thrombotic and bleeding events occurred during the first month of treatment, supporting the clinical rationale for an initial frontloaded standard dose of oral direct factor Xa inhibitor followed by early dose reduction in selected patients. While these findings should be interpreted cautiously given the limited statistical power of the study, generating confirmatory randomized evidence in such a highly vulnerable population may prove particularly challenging.

Our trial incorporated a second-stage randomization within the DXI arm, assigning patients to either apixaban or rivaroxaban. Although the study was not powered for comparisons between these agents, this design provides exploratory data that can be viewed alongside the findings of the COBRRA trial [19], which reported lower bleeding rates with apixaban than rivaroxaban in acute VTE. In VERDICT, point estimates consistently favored apixaban across net clinical benefit, major bleeding, and recurrent VTE outcomes, although confidence intervals were wide and overlapped substantially. Therefore, these findings should be considered hypothesis-generating rather than evidence of differential treatment effects.

Interpretation of the available evidence remains challenging because trials differed substantially in both patient characteristics and anticoagulation strategies. Whereas EINSTEIN and AMPLIFY evaluated standard-dose DXI regimens, HOKUSAI-VTE used dose reduction after initial LMWH therapy, whereas VERDICT evaluated full-intensity oral anticoagulation during the acute phase followed by dose reduction. This distinction may be clinically relevant, as full-dose anticoagulation is required during the period of highest risk of recurrent VTE, whereas subsequent dose reduction may mitigate drug accumulation and bleeding in patients with renal impairment. Despite these differences, incorporation of VERDICT into a meta-analysis did not reveal meaningful heterogeneity, suggesting that our findings are broadly consistent with the existing randomized evidence base and those reported by Van Es et al. [9]. These observations are further supported by contemporary real-world analyses, including those of Cohen et al. [20], which reported comparable outcomes with DXIs and VKA-based regimens in patients with renal impairment.

This study has several limitations. First, the trial used an open-label design; however, central randomization with appropriate concealment and adjudication of all outcomes by an independent blinded committee minimized assessment bias. Second, the trial was prematurely terminated after the inclusion of 200 patients (one quarter of the planned 800 participants). Consequently, the study was underpowered to formally demonstrate noninferiority, though the observed point estimates suggest the objective might have been met had the target sample size been achieved. Finally, the composite endpoint should be interpreted alongside its individual components, as the anticipated clinical benefit was expected to arise primarily from a reduction in major bleeding while maintaining efficacy.

## CONCLUSION

In conclusion, this randomized trial could not demonstrate the non-inferiority for net clinical benefit of early DXI dose reduction strategy compared to standard therapy in patients with VTE and moderate-to-severe renal impairment. Interpretation of these findings is limited by premature trial termination and the resulting lack of statistical power. Nevertheless, VERDICT provides prospective randomized data in a particularly frail and underrepresented patient population for whom high-quality evidence remains scarce.

## Data Availability

The data supporting the findings of this study are available from the corresponding author upon reasonable request.

## Acknowledgments

We sincerely thank all the patients who agreed to participate in this study and thereby contributed to its successful completion. We are also grateful to the data managers (Charly Martin and Laurent Tordella) and clinical research assistants (Caroline Chaudier, Estelle Perrin, and Michaël Pierre) for their invaluable contributions.

The study was designed, conducted, analyzed, and reported in collaboration with the INNOVTE F-CRIN Research Network. We also gratefully acknowledge the contribution of all the clinical research assistants at the participating centers.

## Use of AI

ChatGPT (OpenAI) was used to assist with English language editing. All outputs were reviewed, revised as needed, and approved by the authors, who take full responsibility for the content of the manuscript.

## Author contributions

Patrick Mismetti: conceptualization, methodology, investigation, resources, writing-original draft, writing-review & editing, supervision, project administration, funding acquisition.

Laurent Bertoletti: investigation, resources, writing-review & editing. Antoine Elias: investigation, resources, writing-review & editing.

Carine Assante: data curation, writing-review & editing, project administration. Olivier Sanchez: investigation, resources, writing-review & editing.

Jeannot Schmidt: investigation, resources, writing-review & editing.

Emilie Presles: methodology, software, formal analysis, data curation, writing-review & editing.

Céline Chapelle: formal analysis, writing-original draft, writing-review & editing. Sandrine Accassat: investigation, resources, writing-review & editing.

Francis Couturaud: investigation, resources, writing-review & editing. Isabelle Mahé: investigation, resources, writing-review & editing.

Silvy Laporte: conceptualization, methodology, validation, writing-original draft, writing-review & editing, visualization, supervision, project administration.

## Data sharing

De-identified individual participant data (including text, tables, figures, and appendices) and supporting documents (study protocol, statistical analysis plan, annotated CRFs, and analytic code) will be made available to researchers with a methodologically sound proposal. Requests should be submitted to the corresponding author and will be reviewed and approved by the study steering committee. A signed data sharing access agreement will be required for approved requests. Depending on the weight of VERDICT data in the project, authorship of resulting publications will be discussed.

